# State and trait dimensions of subjective memory concerns: Genetic influences across 30 years

**DOI:** 10.1101/2022.08.08.22278570

**Authors:** Tyler Bell, Asad Beck, Nathan Gillespie, Chandra Reynolds, McKenna E. Williams, Daniel E. Gustavson, Michael J. Lyons, Michael Neale, William S. Kremen, Carol E. Franz

## Abstract

**INTRODUCTION:** Genetic influences on the stability of subjective memory concerns (SMC) may confound its interpretation as a state-related risk indicator.

**METHODS:** We estimated genetic influences on SMC and SMC change from average ages 38 to 67, genetic correlations of SMC with memory and depressive symptoms at average ages 56, 62, and 67, and correlations of SMC with polygenic scores for Alzheimer’s disease and neuroticism, *APOE* genotype, and parental history or dementia.

**RESULTS:** SMC was highly stable over this 30-year period. Heritability was .51. Phenotypically and genetically, SMC was more strongly related to depressive symptoms than to objective memory. It was also correlated with parental history of dementia and polygenic risk for neuroticism.

**DISCUSSION:** SMC’s heritability and stability indicate a substantial trait-like component that is related more to depression and personality than objective memory. Improving it as an indicator of impending cognitive decline or dementia will require isolating its state-related component.

## 1 INTRODUCTION

Approximately 20% of adults report subjective memory concerns (SMC) involving self-reported difficulties and worries about learning and recall [1]. SMC is of significant interest due to its relationship with poorer quality of life [2], poorer mental health [3], and elevated risk of Alzheimer’s Disease (AD) and related dementias [2-6]. Originally, researchers hypothesized that SMC was an state-like measure, capturing new memory decline as one aged [7]. This assumption has been challenged by studies finding weak associations with actual memory performance [8, 9]. In fact, stronger associations are found between SMC and depressive symptoms [10]. With respect to AD and related dementias, the importance of SMC is viewed primarily as an indicator of impending cognitive decline preceding dementia as indicated by the international workgroup that defined the quasi-diagnostic term subjective cognitive decline (SCD) [1]. In potential contradiction with SMC as a marker of imminent decline, a longitudinal study found trait-like stability of SMC (r=.58) assessed at age 40 and 70 [11]. However, this correlation suggests that over 60% of variation in SMC at age 70 is independent of variation at age 40. Furthermore, the trait-like component of SMC warrants further study as this was a relatively small study of only 67 men with data at both timepoints. It is important to better understand what underlies the trait-like component of SMC because any assessment of SMC as a state-related indicator of impending cognitive decline would inevitably include its trait-like component.

In the Adult UK Twin Register, heritability of SMC in adults between 20 and 84 years of age ranged from .37 to .64 [11]. In the Older Australian Twins Study, heritability of SMC was .60 [12]. The remaining variance was attributed to unshared environmental influences. This is surprising because if SMC was due to AD-related cognitive decline, we would not expect SMC to be common in younger adults and to also be heritable.

Instead, frequency and heritability would be thought to be strongest at older ages where SMC arises from genetic AD risk. While cross-sectional, this study suggests there may be a trait-like, heritable component to SMC across age in addition to changes potentially driven by AD. Interestingly, a study of only people with newly occurring subjective cognitive decline reported no heritability [13]. Although SMC may be heritable, we still know nothing about the extent to which the same or different genetic factors may influence SMC at different ages, nor the extent to which shared genetic influences underlie its association with depressive symptoms or objective memory performance. If SMC at later life is related mostly to AD-related changes, we may expect increasing heritability over time. If SMC is long-standing trait reflective of general personality, such as neuroticism, we would expect a heritable trait-like component of SMC across time.

Here we examined genetic and environmental influences on SMC assessed at four timepoints over a 30-year period from ages 38 to 67 as well as SMC’s association with objective memory performance and depressive symptoms. We hypothesized that SMC would be more strongly associated with later SMC and depressive symptoms than with objective memory performance [14], and that shared genetic influences would partially underlie the correlation with depressive symptoms. We expected SMC to be relatively stable as people aged, and we hypothesized that the stable variation would be primarily associated with genetic factors. Biometric twin models assessed for the presence of common and new genetic influences over time. In exploratory analyses, we examined associations of SMC with polygenic risk scores for neuroticism and AD, *APOE* genotype, and parental history of dementia.

## 2 METHODS

### 2.1 Participants

The Vietnam Era Twin Study of Aging (VETSA), is a longitudinal study of cognitive and brain aging and risk for AD in a national U.S. sample of community-dwelling men who served at some time during the Vietnam era (1965-1975) [15-17]. The rate of reported combat exposure was only about 20%. Participants are comparable to American men in their age range on health, education, and lifestyle factors [18].

Methods and initial findings from the VETSA have been discussed elsewhere (http://www.vetsatwins.org/for-researchers/) [15-17]. Participants were first recruited from 2003 to 2007 and were between ages 51 and 60 years. Participants were invited for follow-up assessments approximately 6 and 12 years later. At these waves, average ages were 56 (*n*=520), 62 (*n*=199), and 67 (*n*=1192). The sample size was lower at wave 1 for these analyses because the subjective memory questions were added partway into the study. Details on sample sizes by zygosity are expanded in Table S1.

For ease of interpretation, we will use these average ages to refer to specific waves of assessment. At average age 38, the participants completed questionnaires in the Survey of Health, a study that included a question on SMC [19]. Descriptives of this sample are provided in Table 1. Procedures were approved by the Institutional Review Boards at the University of California, San Diego and Boston University.

**Table 1.**
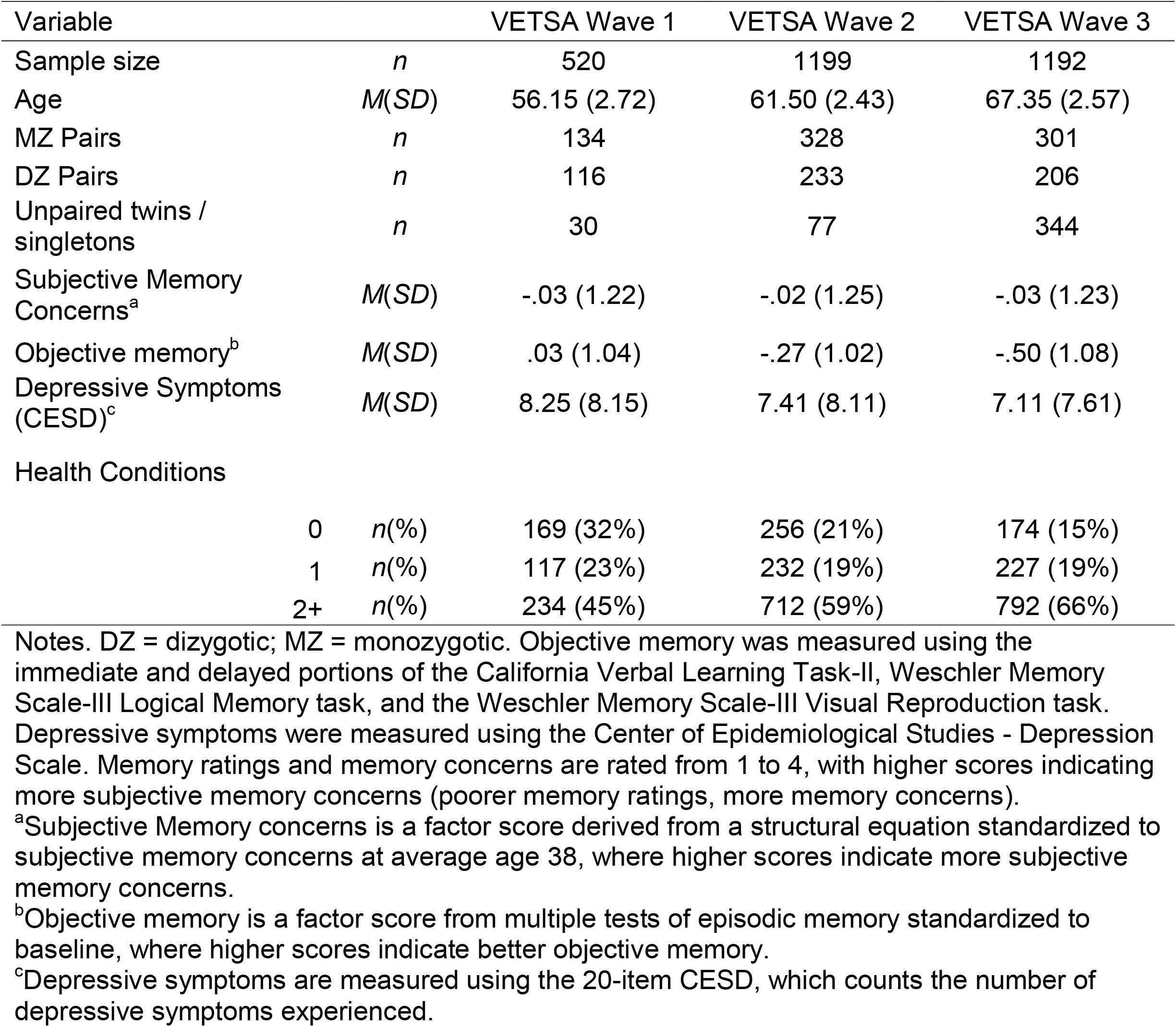
Descriptives of the study sample across waves.

| Variable |  | VETSA Wave 1 | VETSA Wave 2 | VETSA Wave 3 |
| --- | --- | --- | --- | --- |
| Sample size | *n* | 520 | 1199 | 1192 |
| Age | *M(SD)* | 56.15 (2.72) | 61.50 (2.43) | 67.35 (2.57) |
| MZ Pairs | *n* | 134 | 328 | 301 |
| DZ Pairs | *n* | 116 | 233 | 206 |
| Unpaired twins /<br>singletons | *n* | 30 | 77 | 344 |
| Subjective Memory<br>Concerns<sup>a</sup> | *M(SD)* | -.03 (1.22) | -.02 (1.25) | -.03 (1.23) |
| Objective memory<sup>b</sup> | *M(SD)* | .03 (1.04) | -.27 (1.02) | -.50 (1.08) |
| Depressive Symptoms<br>(CESD)<sup>c</sup> | *M(SD)* | 8.25 (8.15) | 7.41 (8.11) | 7.11 (7.61) |
| Health Conditions |  |  |  |  |
|  | 0 *n*(%) | 169 (32%) | 256 (21%) | 174 (15%) |
|  | 1 *n*(%) | 117 (23%) | 232 (19%) | 227 (19%) |
|  | 2+ *n*(%) | 234 (45%) | 712 (59%) | 792 (66%) |

Notes. DZ = dizygotic; MZ = monozygotic. Objective memory was measured using the immediate and delayed portions of the California Verbal Learning Task-II, Weschler Memory Scale-III Logical Memory task, and the Weschler Memory Scale-III Visual Reproduction task. Depressive symptoms were measured using the Center of Epidemiological Studies - Depression Scale. Memory ratings and memory concerns are rated from 1 to 4, with higher scores indicating more subjective memory concerns (poorer memory ratings, more memory concerns).
<sup>a</sup>Subjective Memory concerns is a factor score derived from a structural equation standardized to subjective memory concerns at average age 38, where higher scores indicate more subjective memory concerns.
<sup>b</sup>Objective memory is a factor score from multiple tests of episodic memory standardized to baseline, where higher scores indicate better objective memory.
<sup>c</sup>Depressive symptoms are measured using the 20-item CESD, which counts the number of depressive symptoms experienced.

### 2.2. Subjective memory concerns

At age 38, SMC was scored using a single item, which was converted into a continuous threshold liability score and standardized. At ages 56, 62, and 67, SMC was scored using 2 items, which were combined into a latent factor score fixing the factor loadings at 0.50 with standardized mean and variance of 0 and 1 respectively. SMC was centered at age 38. As such, in subsequent waves, a score of 0 indicates no change, higher scores indicate greater SMC, and lower scores indicate less SMC after age 38. Table S2 shows question items for each wave.

### 2.3. Objective memory performance

At average ages 56, 62, and 67, objective memory performance was measured using the immediate and delayed recall portions of the California Verbal Learning Test-II [20], and the Weschler Memory Scale-III Logical Memory and Visual Reproduction subtests [21]. Our previous confirmatory factor analysis show that items on these tests can be best explained by a highly heritable common latent factor [22]. Each subject’s latent factor score was adjusted for practice effects as previously described elsewhere [23]. Factor scores at average ages 62 and 67 were fitted in reference to objective memory performance at average age 56. As such, 0 represents performance similar to performance at average age 56, >0 indicates performance improvement from age 56, and <0 represents performance decline from average age 56. There were no objective memory data available from age 38.

### 2.4 Additional measures for exploratory analyses

We also examined polygenic risk scores (PRSs) for AD [24, 25] and neuroticism [26], which were adjusted for five principal components of ancestry. Methods for deriving PRSs are described in detail elsewhere [26, 27]. We also examined associations with *APOE* genotype (ε4+ vs. ε4-) and parental history of dementia. Participants were asked, “Did your biological mother/father ever have a serious memory difficulty referred to as senility, Alzheimer’s disease, or other kind of dementia”. Discordant twin pairs i.e., where twins disagreed regarding parental history were coded as missing to avoid ambiguity.

### 2.5. Physical health conditions

The number of health conditions was measured using an index of the history of hypertension, heart attack, heart failure, stroke, peripheral vascular disease, thrombolysis, angina, diabetes, bronchitis, asthma, cancer, osteoarthritis, rheumatoid arthritis, or cirrhosis reported during the medical interview at each study wave (range from 0 to 14). These conditions were chosen as they are listed in the Charlson Index, a validated index of major medical conditions [28].

### 2.6. Statistical analysis

#### 2.6.1 Phenotypic Analyses

We first used linear mixed models to characterize the change in SMC at each wave. SMC was regressed onto age as our longitudinal time variable. Random intercepts were used to adjust for the non-independence of SMC reported within individuals and within twin pairs. Scatterplots were used to visualize trends over time.

Next, we tested multivariate relationships using structural equation models in Mplus version 8.4. We used latent variables to provide composites for SMC at ages 56, 62, and 67. SMC at age 38 was based on only one item and left as an observed variable. The structural equation model included: 1) autoregressive (unidirectional) paths from observations within the same measure over time; 2) bidirectional paths that were set at average age 56, 62, and 67 between SMC and objective memory, SMC and depressive symptoms, and objective memory and depressive symptoms; and 3) lagged paths from all variables to all variables at subsequent waves. The model was adjusted for age and number of health conditions at each wave. We used the TYPE=COMPLEX option to adjust standard errors for dependent data between twins in the same pair. The model incorporated general cognitive ability at average age 20 and SMC at age 38 as predictors of SMC, objective memory, and depressive symptoms at ages 56, 62, and 67.

#### 2.6.2 Biometric Twin Analyses

The OpenMx2.9.9.1 software package [29] in R3.4.1 [30] was used to fit univariate and multivariate biometric genetic twin models [31] estimating the relative contribution of genetic and environmental influences within and between the SMC measures [32, 33]. Types of multivariate models are shown in Figure 1. We then created extended biometric models of SMC with objective memory and depressive symptoms at ages 56, 62, and 67 (waves when all variables were assessed) to examine genetic and environmental correlations of SMC with objective memory and depressive symptoms. All code is provided online (https://github.com/trbellucsd/SubjectiveMemory). More details about analyses are listed in the Supplemental Information.

**Figure 1.**
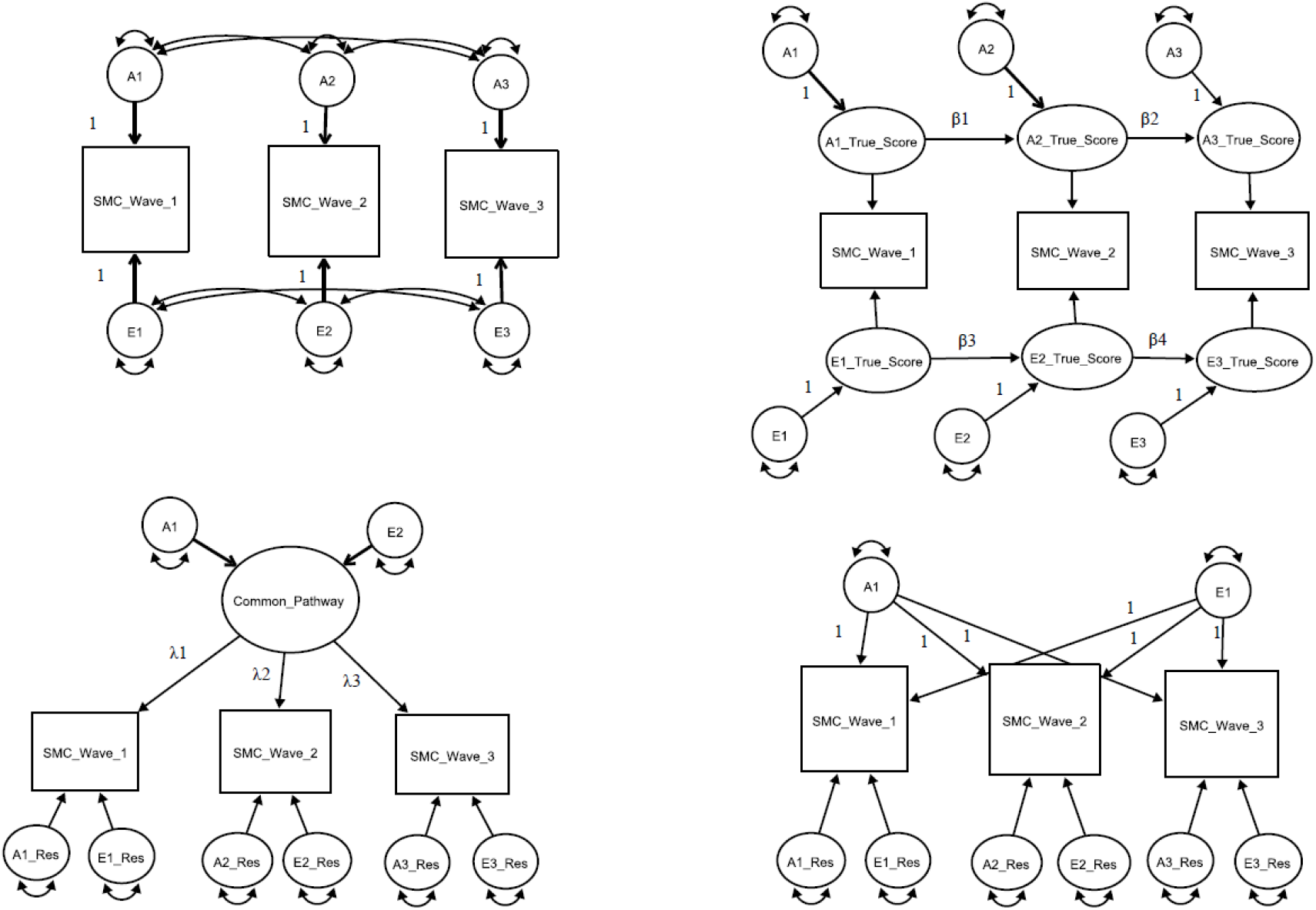
Types of multivariate biometric genetic twin models. Note. Multivariate correlated factors (A) and competing hypothetical models to explain the sources of variance-covariance between subjective memory concerns (SMC) at each assessment wave. Competing models include (B) the auto-regression, (C) common pathway, and (D) independent pathway models. For brevity, only latent additive genetic (A1–3) and non-shared environmental (E1–3) factors are shown. (A) The multivariate correlated factor model estimates the size of the latent genetic and environmental variances and covariances (double-headed arrows). It is atheoretical and makes no prediction about the nature of change in SMC over time. (B) In the autoregression model, the time-specific genetic (variance of A1−3) and environmental (variance of A1−3) variance components or “innovations” for each genetic (A1–3) and environmental (E1–3) latent factor true score are estimated along with each variable’s residual or error variance (residual of E1−E3). Also estimated are the autoregression or causal coefficients (β) from one latent true score to the next. (C) In the common pathway model, the genetic (A1) and environmental (E1) variance components for the common pathway, the factor loadings (paths not set to 1 from latent factor), and latent genetic and environmental residuals are estimated. (D) Finally, in the independent pathway model, genetic and environmental variance components are estimated independently with their factor loadings, and latent genetic and environmental residuals.

#### 2.6.3 Exploratory analyses

In exploratory analyses, we examined polychoric correlations between SMC at ages 38, 56, 62, and 67 and polygenic risk scores for AD and neuroticism, as well as *APOE* genotype (ε4+ vs. ε4-) and parental history of dementia. Polychoric correlations were calculated using the OpenMx_2.9.9.1_ software package [29] in R_3.4.1_ [30] and were adjusted for the pairing of continuous and categorical variables by applying threshold liability estimation for ordinal and binary variables. Significance levels for set at *p* < .05. For the biometric models, we present 95% percent confidence intervals (CI), in which an interval not including zero is considered significant.

## 3 RESULTS

### 3.1 Phenotypic analyses

There was no significant change in SMC over time in the age-based linear mixed models (*p* = .629). We then moved into wave-based SEM analyses to examine SMC over time and their phenotypic, multivariate associations with depressive symptoms and objective memory as shown in Figure 2 (see Table S3 for bivariate correlations).

**Figure 2.**
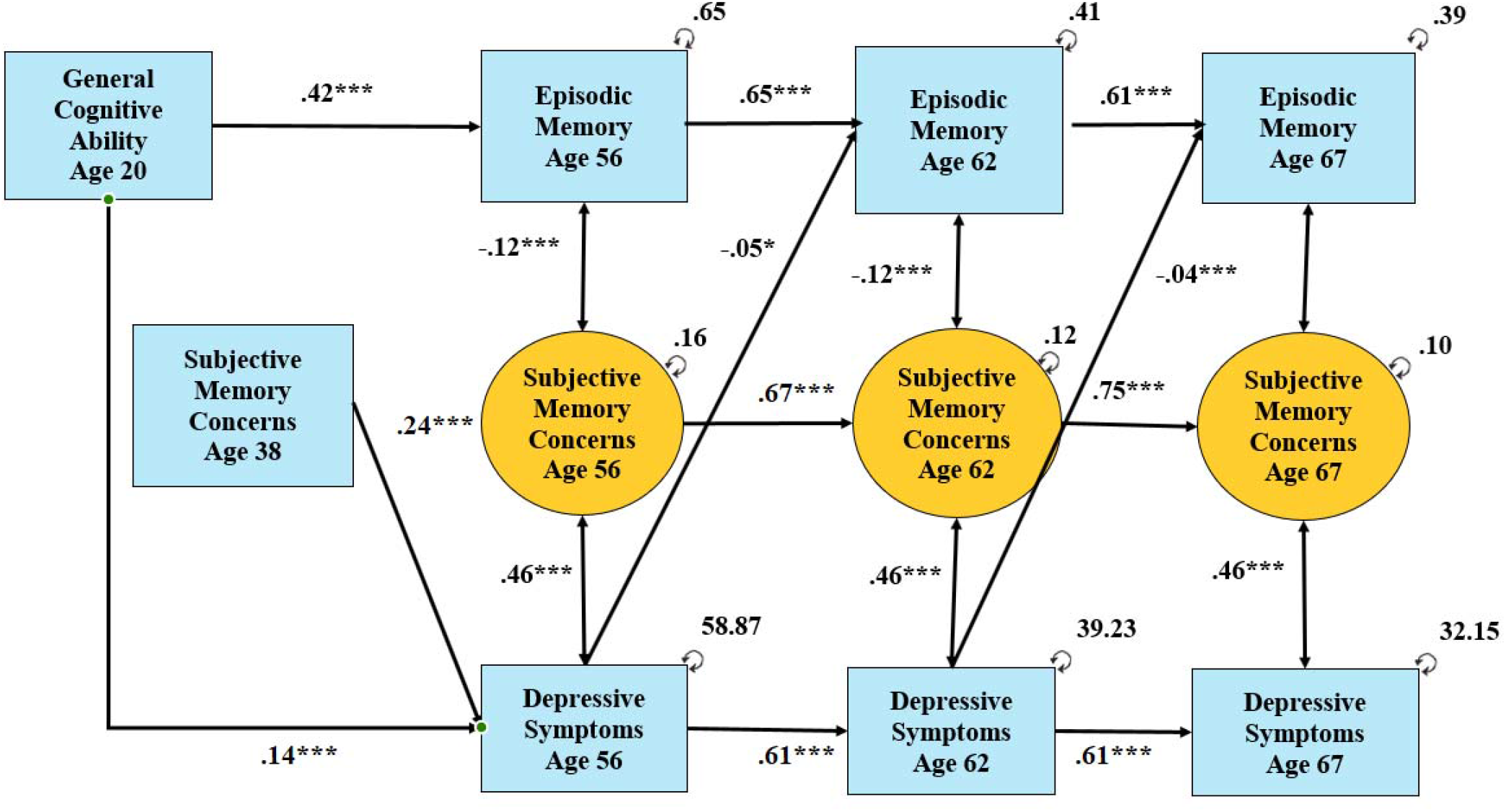
Structural equation model of phenotypic associations between subjective memory concerns, objective memory, and depressive symptoms. Note. This structural equation model is additionally adjusted for age, the number of health conditions, and clustering of twin pairs. For readability, only significant paths (*p*s < .05) are shown. The model included a test for all possible bidirectional and lagged paths between variables. The labeled ages stand for the average age of the sample at the wave when the variable was collected. Note that subjective memory concerns at age 38 were an observed variable (as it only included one item) compared to subjective memory concerns at ages 56, 62, and 67, which are latent variables (as there were two items measured at these waves).

Regarding the earliest measured variables, SMC at age 38 was related to SMC at age 56 (β=.24, *p*<.001). Greater general cognitive ability at age 20 was related to better objective memory at age 56 (β=.42, *p*<.001). At ages 56, 62, and 67, all autoregressive paths were significant and largest for SMC (βs=.67 to .75, *p*s<.001), objective memory (βs=.65 and .61, *p*s<.001), and depressive symptoms (βs=.61, *p*s<.001), respectively. Significant concurrent associations could also be fixed without significant deterioration in -2LL (*p*s>.05). Specifically, SMC was associated with concurrent lower objective memory (βs=-.12, *p*s<.001) and more depressive symptoms (βs=.61, *p*s<.001). Objective memory was not associated with current depressive symptoms (*p*=.102). Looking at lagged associations, depressive symptoms were associated with worse objective memory at the subsequent waves (βs=-.05 and -.04, *p*s<.05), but not the reverse. No other significant lagged associations were noted.

### 3.2 Biometric twin analyses

Next, we ran a correlated liabilities model to obtain multivariate heritability estimates for SMC. See Table 2 for MZ/DZ correlations. Univariate and multivariate estimates of heritability are presented in Supplemental Tables S3 and S4, respectively. Derived from the multivariate correlated factors model, SMC at age 38 was genetically correlated with SMC at ages 56 (*r*_A_=.63), 62 (*r*_A_=.56), and 67 (*r*_A_=.49); genetic correlations between SMC at ages 56, 62, and 67 were high (*r*_A_s=.78 to .90) (see Table 2).

**Table 2.** Within Pair, Phenotypic, and Genetic Correlations of Subjective Memory Concerns (SMC).

| Variables | MZ | DZ | Phenotypic (below diagonal) and<br>Genetic Correlations (above diagonal) |  |  |  |
| --- | --- | --- | --- | --- | --- | --- |
|  |  |  | 1 | 2 | 3 | 4 |
| 1. SMC Age 38 | **0.21** | **0.18** | - | **.63** | **.56** | **.49** |
| 2. SMC Age 56 | **0.34** | **0.12** | **.27** | - | **.90** | **.78** |
| 3. SMC Age 62 | **0.29** | **0.11** | **.19** | **.52** | - | **.87** |
| 4. SMC Age 67 | **0.38** | 0.05 | **.25** | **.47** | **.59** | - |

Note. MZ = monozygotic twins; DZ = dizygotic twins; SMC = subjective memory concerns.
Significant *p*-values are bolded.

Looking at multivariate models, the best fitting ACE model was the common pathway model compared to the autoregressive or independent pathway models (Table S5, for which the C component was able to be dropped without significant deterioration in model fit (Table S6). As shown in Figure 3, the latent liability of SMC was heritable (heritability =.51, 95% CI:.40; .61). Similarly, the common pathway models fit better than autoregressive or independent pathway models for objective memory and depressive symptoms (Table S7). Furthermore, in biometric twin models, the C component was able to be dropped without significant deterioration in model fit for objective memory and depressive symptoms (Table S8), showing significant heritability for latent liabilities for objective memory (heritability =.74, 95% CI: .68; .79) and depressive symptoms (heritability=.40, 95% CI: .14; .66).

**Figure 3.**
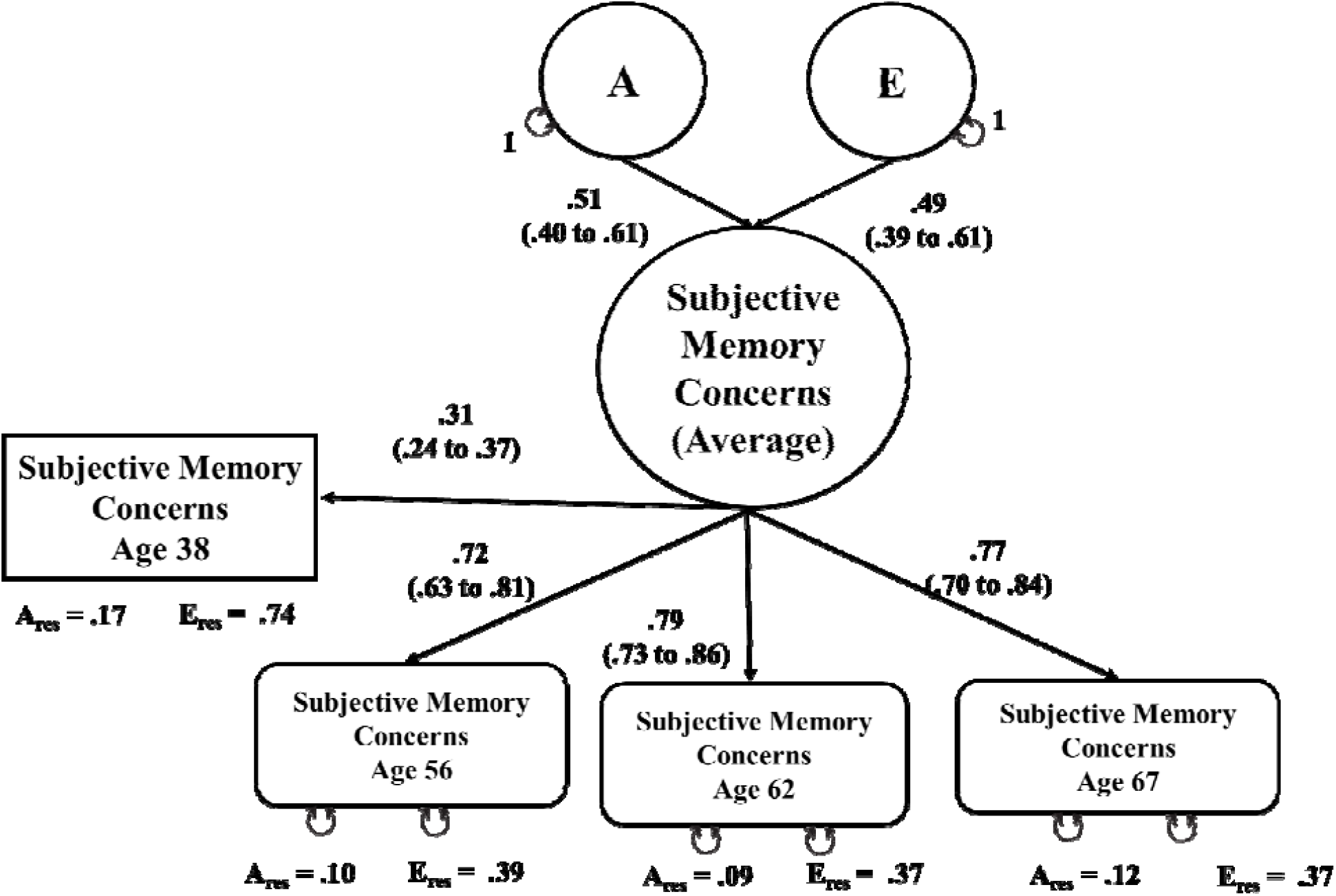
Common pathway model of subjective memory concerns. Note. A = additive genetic influence (heritability); A res. = residual for genetic influences (A); E = unshared environmental influences; E res. = residual for unshared environmental influences (E). Subjective memory concerns at age 38 is an observed measure from one item described in the measures. Subjective memory concerns at age 56, 62, and 67 are factor scores from two items from the phenotypic SEM model.

Extended biometric twin models showed show significant correlations of SMC with the latent liabilities of objective memory and depressive symptoms, in Figures 4 and 5 respectively, and in the expected directions. Genetic correlations (*r*_A_) were small between latent liabilities for SMC and objective memory (*r*_A_ [genetic correlation]=-.13, 95% CI:-.21; -.05) and moderate between latent liabilities for SMC and depressive symptoms (*r*_A_=.29, 95% CI: .20; .36). Unshared environmental correlations (*r*_E_) followed a similar pattern for SMC with objective memory (*r*_E_=-.08, 95% CI:-.14; -.03) and SMC with depressive symptoms (*r*_E_= .34; 95% CI:.26; .41)

**Figure 4.**
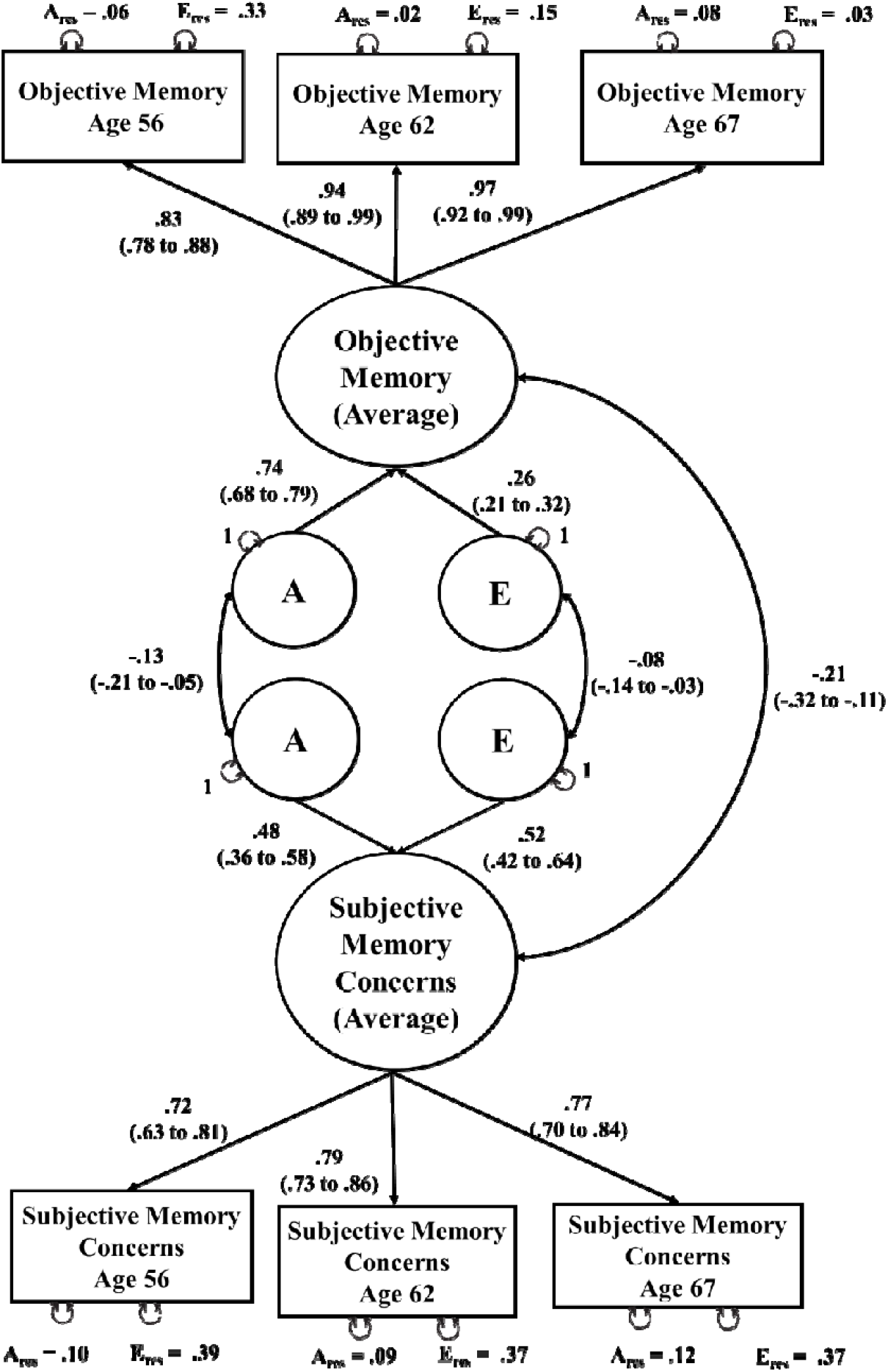
Common pathway model showing associations between subjective memory concerns and objective memory. Note. A = additive genetic influence (heritability); A res. = residual for genetic influences (A); E = unshared environmental influences; E res. = residual for unshared environmental influences (E).

**Figure 5.**
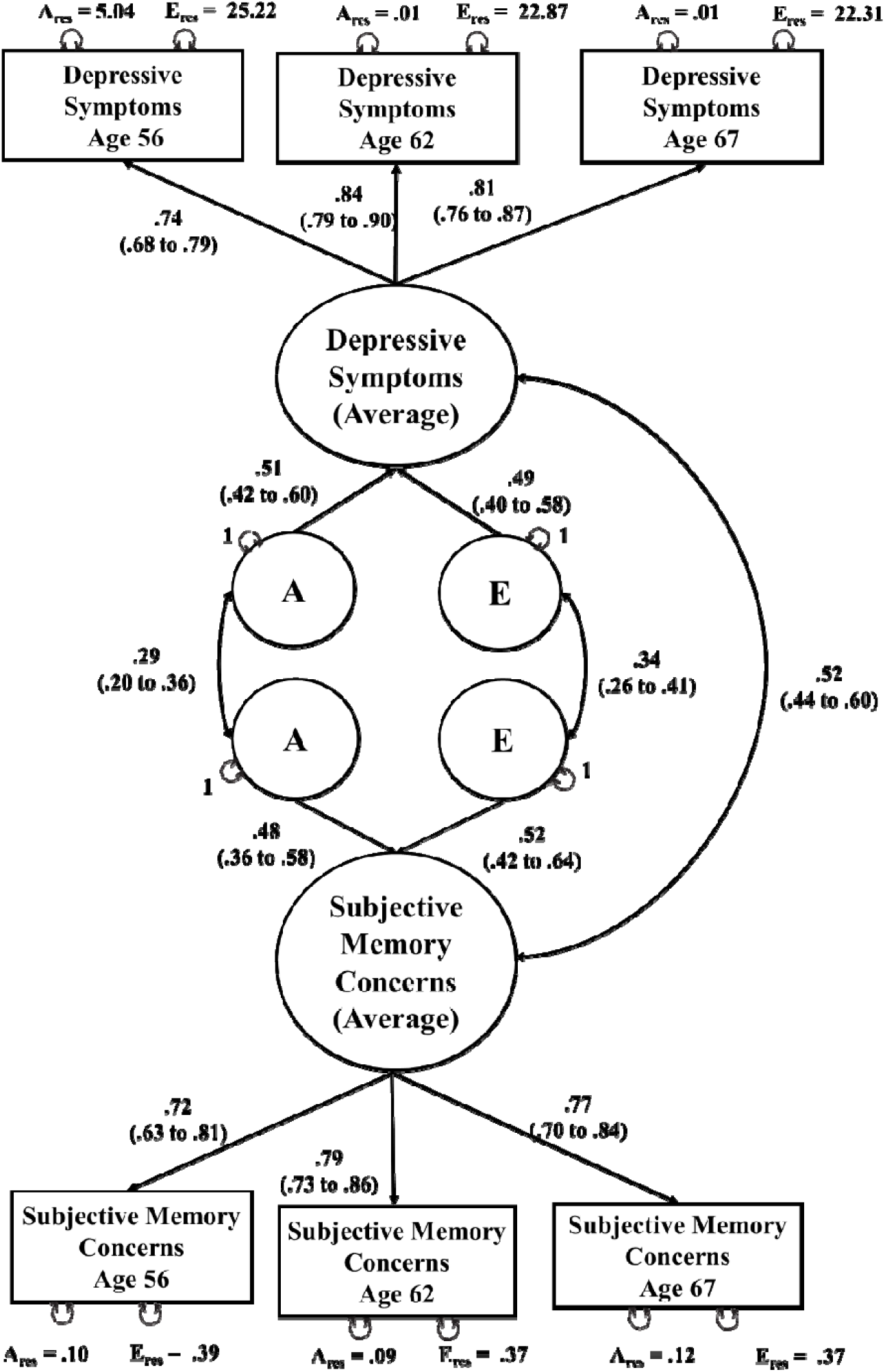
Common pathway model showing the association between subjective memory concerns and depressive symptoms. Note. A = additive genetic influence (heritability); ; A res. = residual for genetic influences (A); E = unshared environmental influences; E res. = residual for unshared environmental influences (E).

### Exploratory Analyses

We also explored how average SMC related to other genetic and familial risk factors. The latent liability of SMC was not significantly related to the polygenic risk score for AD or to *APOE* genotype. However, the latent liability SMC was significantly positively associated with the polygenic risk score for neuroticism (*r*=.08, *p*=.018) and with parental history of dementia (*r*=.12, *p*<.001). Correlations at ages 38, 56, 62, and 67 are also shown in Table 3.

**Table 3.**
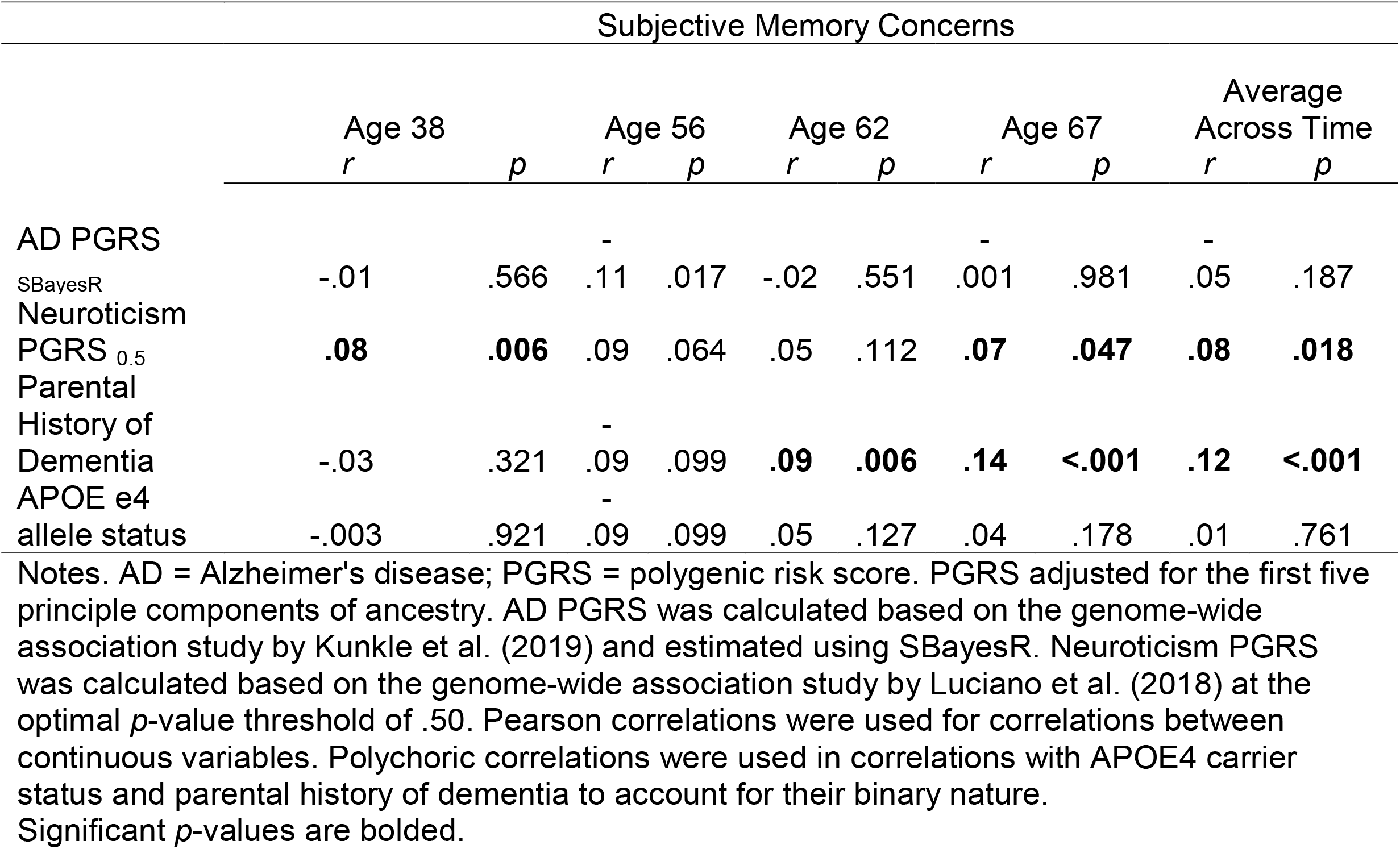
Exploratory associations between subjective memory concerns and genetic risk factors.

## 4 DISCUSSION

We examined SMC over a 30-year period and its relationship to objective memory performance and depressive symptoms in a group of community-dwelling men. SMC did not show a general increase as objective memory declined across 3 decades and were more reflective of individual differences in depressive symptoms than memory performance. The best-fitting model supported a single underlying latent phenotype that is responsible for the covariation among the SMC measures at different ages. Genetic influences accounted for about half (.51) of the variance in the SMC latent phenotype.

Biometric models also showed a moderate amount of shared genetic influences between SMC and depressive symptoms but only a small amount of shared genetic influences between SMC and objective memory performance. Average SMC was additionally associated with polygenic risk for neuroticism and with parental history of dementia.

In several studies of older adults, SMC has been shown to predict cognitive decline and dementia within 3 to 4 years [7, 34], but others have shown similar rates of future cognitive decline in older adults with and without SMC [14, 35, 36]. Here we found evidence for a weak concurrent association between SMC and level of actual objective memory performance, which has been shown in a sample much older than ours as well [37], but no evidence of SMC predicting future memory performance. There is also evidence that associations between SMC and objective memory are due to general differences at baseline rather than correlated changes across older adulthood [8]. Here, we expand this to show that this seems true when at looking at general differences from later midlife as well.

Taken together, our results are consistent with the idea that SMC involves a trait-like component from very early midlife to early old age. Despite significant declines in objective memory performance across assessment periods in the present study, SMC remained generally stable. The high genetic correlations of SMC over a 30-year period suggest a relatively stable trait-like dimension. Moreover, even SMC at age 38—long before a time when age-related memory decline is thought to occur—was significantly correlated with SMC 20 and more years later [38]. These results build substantially on the one prior small sample study with only two timepoints [11], while also able to estimate genetic and environmental influences due to the twin nature of the data.

Consistent with prior studies [14, 15], we showed that about half of the variance in SMC is accounted for by genetic influences. We also demonstrated that a common latent phenotype could best account for SMC across all four timepoints, and the latent phenotype of SMC over time was primarily due to a trait-like genetic factor.

In contrast to objective memory performance, SMC was more strongly related to depressive symptoms, as suggested in prior studies [3, 10, 39, 40]. As a twin study, we were able to examine these relationships beyond the phenotypic level. Our analysis showed a significant genetic correlation between SMC and depressive symptoms, indicating some shared genetic influences. Taken together with the genetic correlation between SMC and depressive symptoms, we hypothesized that SMC might be genetically related to another trait-like characteristic, namely, personality. Given strong genetic links between depression and anxiety [41], we thus examined the association between SMC and a polygenic risk score for neuroticism. Neuroticism is defined as the tendency to negatively evaluate one’s world and self, leading to distress, fear, and avoidance, likely based on high levels of physiological reactivity to stimuli [42]. Like SMC, neuroticism is heritable, stable across adulthood, and related to general levels of depressive symptoms [43]. As would be expected given the substantial genetic overlap between anxiety and depression, individuals with higher SMC were also at higher genetic risk for neuroticism based on the polygenic score. Genes involved in neuroticism have been implicated in the development of brain structures for attention and learning (e.g., anterior cingulate cortex, amygdala, medial temporal lobe, and hippocampus) [44-47], which may be involved in attentional biases toward worrying and distress.

SMC was not related to either the AD polygenetic risk score or *APOE* genotype, but it was related to parental history of dementia, perhaps suggesting greater links to general dementia risk. A similar association has been recently found in the Health and Retirement Study, the largest analysis of its kind [48]. It could be that having a parent with dementia increases one’s trait levels of SMC, or that dementia-related genes not captured by the AD polygenic risk score underlie the association between SMC and later dementia risk. Future work is needed and SMC measures accounting for the strong trait-like component may improve associations between SMC and genetic AD risk.

Lastly, about half of the variation in SMC was due to unshared rather than shared environmental influences. Such a finding suggests an important role of unique life experiences in how people form their beliefs about subjective memory early on.

Negative life experiences can lead to reduced beliefs about one’s abilities, which may generalize to evaluations of their cognitive abilities. This can include experiences with major health concerns and social stressors in adolescence and young adulthood, which may leave one feeling more worried about one’s abilities [49]. It may also include evaluations of success in interpersonal relationships, academic performance, and sports [50]. More work will be needed to discern early life events impacting longstanding perceptions of subjective memory.

Our study had limitations to consider. Regarding external validity, our sample may not be generalizable to other groups. For one, our sample was 100% men, limiting generalizability to women. Gender and sex differences have been noted in the association between SMC and objective memory. For example, females who report SMC are more likely to have memory decline, unlike men who showed no such association [51]. Our sample also primarily identified as non-Hispanic White, similar to previous literature on this topic [8]. More diverse samples may show relationships between SMC and memory decline, especially older Latino adults [52, 53]. Our sample came from a community-dwelling population, which may not generalize to individuals seen in clinical settings.

SMC in older adults lowers quality of life, is associated with negative mental health outcomes and elevated the risk for dementia [34]. As such, the nature of SMC is interest for more fully understanding the etiology of these relationships. SMC was originally conceptualized as a state-related factor indirectly capturing memory decline. However, in the present study we found that SMC is a heritable, trait-like phenomenon in older men that was more strongly related to depressive symptoms than to objective memory at both the phenotypic and genetic levels. Furthermore, SMC at follow-up assessments were not related to earlier changes in depressive symptoms or objective memory performance. Consideration of longstanding memory concerns may help researchers and clinicians better detect reports related to actual memory decline. Put another way, it will be important to isolate the state-related component of SMC to improve its utility as a risk indicator of impending cognitive decline or dementia.

## Data Availability

Data is available upon request and approval of a data authorization agreement.

## Conflict of Interest Declaration

The authors declare the absence of known competing financial or personal relationships that could have influenced the work reported in this paper.

## Financial Support

This work was supported by the National Institute on Aging at the National Institutes of Health grant numbers R01s AG050595 (WSK MJL CEF), AG022381 (WSK), AG037985, AG062483, and P01 AG055367.

## Notes

### Competing Interest Statement

The authors have declared no competing interest.

### Author Declarations

Ethical approval was granted by the Institutional Review Board of the University of California San Diego University.

